# Antiretroviral therapy associated lipodystrophy; learning from the past

**DOI:** 10.1101/2021.04.18.21255695

**Authors:** Senai Goitom Sereke, Semhar Eyob Berhe, Felix Bongomin

## Abstract

**Background:** With the introduction of effective antiretroviral therapy (ART), descriptions of body shape abnormalities, such as central fat accumulation and peripheral fat loss emerged among persons living with human immunodeficiency virus (HIV). We aimed to determine the prevalence of lipodystrophy and associated risk factors among patients on ART at Orotta National Referral Hospital (ONRH), Asmara, Eritrea.

**Methods:** A single center, retrospective study was conducted at the ONRH, reviewing records of HIV-infected patients commenced on ART between January 2007 and December 2012.

**Results:** Records of 250 eligible patients were reviewed. Most were female (59.2%, n=148) with a median age of 35 (IQR-20-63) years. Forty-three (17.2%) participants had body fat abnormalities. 42 (97.6%) had lipoatrophy and 1 (2.4%) buffalo hump. Of the 43 patients with lipodystrophy 34 (79%) were on Stavudine (d4T)/Lamivudine (3TC)/Nevirapine (NVP) regimen, 6 (14%) on Zidovudine (AZT)/3TC/NVP, 2 (4.7%) on d4T/3TC/Efavirenz (EFV) and 1(2.3%) on AZT, 3TC, EFV. EFV-based regimen was significantly associated with lipodystrophy (p< 0.01).

**Conclusion:** We report a high prevalence of lipodystrophy. Four drug regimens were incriminated in the development of lipodystrophy. EFV-based regimen was significantly associated with the lipodystrophy.

## Background

The human immunodeficiency virus (HIV) pandemic has spread to every country in the world and has claimed over 33 million lives worldwide (1). The identification of a cytopathic retrovirus in 1983 and development of a diagnostic serologic test for HIV-1 in 1985 played a major role in developing improvements in the diagnosis of the disease condition (2). Moreover, management was dramatically altered with the introduction of antiretroviral drugs in 1987 and in 1996 it was revolutionized by combination treatment, known as highly active antiretroviral therapy (HAART) (3). In the initial years, following the introduction of HAART, mortality, AIDS, AIDS-defining disease, and hospitalizations significantly decreased by 60 to 80 percent (4).

Antiretroviral therapy (ART) strategy uses combination of several antiretroviral drugs from different classes and mechanisms of action to treatment of HIV infection (5). The primary goal of ART is to increase disease-free survival through suppression of viral replication and improvement in immunologic function (6).

HIV-associated lipodystrophy typically refers to changes in fat distribution that are often associated with metabolic abnormalities, including dyslipidemia and insulin resistance (7). Patients with lipoatrophy have loss of subcutaneous fat mostly noticed in their limbs, face, and/or buttocks areas, and patients with fat accumulation have gain of visceral fat in the abdomen, dorsocervical fat pad enlargement (buffalo hump) and breast enlargement (8,9). Different studies suggest that use of thymidine analogue nucleoside reverse transcriptase inhibitors (stavudine and, to a lesser extent, zidovudine) is strongly associated with the development of lipoatrophy (9,10). Another observational data also suggested that use of protease inhibitors with nucleoside reverse transcriptase inhibitors may accelerate the rate of fat loss (11).

There are multiple reasons to evaluate the body fat abnormalities in patients on ART. Patients are often distressed by the physical changes in their appearance, especially if they develop facial lipoatrophy. These morphologic abnormalities can have a significant impact on self-esteem (12). Lipodystrophy, or fear of developing it, may impact adherence to ART or the willingness to initiate such therapy. Correction of lipodystrophy may improve quality of life (13). Thus, a better understanding of drug side effects and potential risk factors for developing body fat abnormalities will affect the overall picture of patient care and management. Identifying the drug regimen that is incriminated for the disorder and risk factors associated with are very crucial. Therefore, our study aimed at retrospectively reviewing the prevalence of body fat abnormalities in patients on ART at the Orotta National Referral Hospital (ONRH), a large tertiary hospital in Eritrea. The information that is generated from this study will be used to inform clinical practice and policy.

## Methods

### Study design and setting

This was a retrospective, descriptive and single-centered study conducted at the Orotta National Referral Hospital, Asmara, Eritrea. Orotta National Referral Hospital is a tertiary hospital, located at Senita 2km from Asmara city center. In this hospital HIV clinic and ART center is managed by 4 counselors, one data manager and one Internist. In 2005, Orotta ART center was one of the five centers where ART provision and patients follow up started. It provided more than 55 percent of the whole ART follow ups in the country. In this center from January 2007 to December 2011, there were 1868 ELISA positive patients. Out of this 1129 patients were started on ART over similar duration (HIV clinic register Dec 2015).

### Study population

All patients who were diagnosed with HIV based on national algorithm and were on ART treatment in the period between January 2010 and December 2015 were randomly recruited. Patients with missing follows up and missing data were excluded.

### Sample Size

The sample size was determined using computer Programme for epidemiologist (PEPI), version 3.01, described by Armitage and Berry, and 250 participants were determined as the sample size of the study. This was 22 percent of the total population on ART.

### Study procedure

Initially patients’ name was put in alphabetical order. And number was given to the names. This study was designed to generate information from patients’ clinical follow up cards documented by a clinician at every visit. Then we selected computer’s random numbers. The data collecting tool was a self-prepared form. After collecting the data, data entry was done in Microsoft Excel and is given to a professional biostatistician for analysis of the numerical data in SPSS (version 21.0, 2013) and inferring proportions in a whole from these in representative sample.

### Statistical analysis

Categorical data was summarized as frequency and percentages and numerical data as mean ± standard deviations or median (range). Demographic, immunologic, clinical, laboratory, and drug treatment factors were assessed in startatified and multivariate analyses for their relationship to the presence and severity of fat accumulation and atrophy. For specific drug regimens confidence interval was done. P<0.05 was considered statistically significant.

## Results

We reviewed records of 250. The median(range) age of the patients was 35(20-63) years and 148(59.2%) were women. The baseline median(range) weight for all the patients was 51.7(34-82) kg.

Most of the patients started taking ART in the year 2005 (n=71, 28.4%), 2006 (n=75,30%), and 2007 (n=59, 23.6%). Out of 247 with recorded treatment regimen, 157(62.6%) had a treatment regimen, the majority, 157 (62.6%) were on d4T/3TC/NVP and also for those(n=232) with WHO clinical stages, 129(55.6%) were in WHO clinical stage III. Majority of the patients (n=210, 84%) had excellent self-reported ART compliance. Baseline median(range) CD4 count was 151(7-993). The median (range) WBC count was 4.1(1-32). The median(range) of hemoglobin (Hb), platelets (PLT), Aspartate Aminotransferase (AST), Alanine Aminotransferase (ALT), Alkaline phosphatase (ALK.PH), Albumin (ALB), Bilirubin (BIL), Blood-urea nitrogen (BUN), and Creatinine (Cr) were 12.4(8-18), 215(37-677), 33.5(10-284), 24 (3-366), 75(7-919), 3.6(0-5), 0.4(0-3), 9(1-48), and 0.7(0-2) respectively (**Table 1**).

**Table 1:** Sociodemographic and clinical characteristics of the participants.

| Characteristics | Frequency/Median/Mean | Percent/SD/Range |
| --- | --- | --- |
| **Age** | 35 | 20-63 |
| **sex** |  |  |
| Female | 148 | 59.2 |
| Male | 102 | 40.8 |
| **Body weight at baseline** | 51.25 | 34-81.5 |
| **Year when ART was started** |  |  |
| 2005 | 71 | 28.4 |
| 2006 | 75 | 30 |
| 2007 | 59 | 23.6 |
| 2008 | 28 | 11.2 |
| 2009 | 17 | 6.8 |
| **Regimen(n=247)** |  |  |
| ABC/3TC/EFV | 1 | 0.4 |
| ABC/DDI/EFV | 1 | 0.4 |
| AZT/3TC/ABC | 1 | 0.4 |
| AZT/3TC/EFV | 26 | 10.5 |
| AZT/3TC/NVP | 33 | 13.4 |
| D4T/3TC/EFV | 28 | 11.3 |
| D4T/3TC/NVP | 157 | 62.6 |
| **Regimen 1(n=247)** |  |  |
| ABC based | 2 | 0.8 |
| D4T based | 60 | 24.3 |
| AZT based | 185 | 74.9 |
| **Regimen 2 (n=247)** |  |  |
| EFV based | 56 | 22.8 |
| NVP based | 190 | 76.9 |
| Triple nukes | 1 | 0.4 |
| **WHO clinical stage** | n=232 |  |
| I | 50 | 21.6 |
| II | 43 | 18.5 |
| III | 129 | 55.6 |
| IV | 10 | 4.3 |
| **CD4( baseline)** | 151 | 7-993 |
| **Blood count** |  |  |
| WBC | 4.1 | 0.5-320 |
| HGB | 12.4 | 7.6-18.3 |
| PLT | 215 | 116-245 |
| AST | 33.5 | 10-284 |
| ALT | 24 | 3-366 |
| ALK.PH | 75 | 7.2-919 |
| ALB | 3.60 | 0.1-2.7 |
| BIL | 0.4 | 0.1-2.7 |
| BUN | 9 | 0.6-48 |
| CR | 0.7 | 0.1-2.3 |
| **Drug compliance(n=239)** |  |  |
| Poor | 12 | 5.02 |
| Good | 4 | 1.7 |
| Excellent | 210 | 87.9 |
| Drop out | 9 | 3.8 |
| Drop out (defaulter) | 4 | 1.7 |
| **Regimen change** |  |  |
| Yes | 82 | 32.8 |
| No | 168 | 67.2 |

The median CD4 count of the patients increased steadily from 194 cell/microliter at the baseline to 351 cells/microliter at visit 6 (p<0.0001) (Figure:1). On the other hand, the median weight of the participants increased from 51.25kgs at baseline to 55.0kgs at visit 2 (P-0.03) and then remained relatively constant till visit 6 (p=0.24) (Figure: 2).

**Figure 1:**
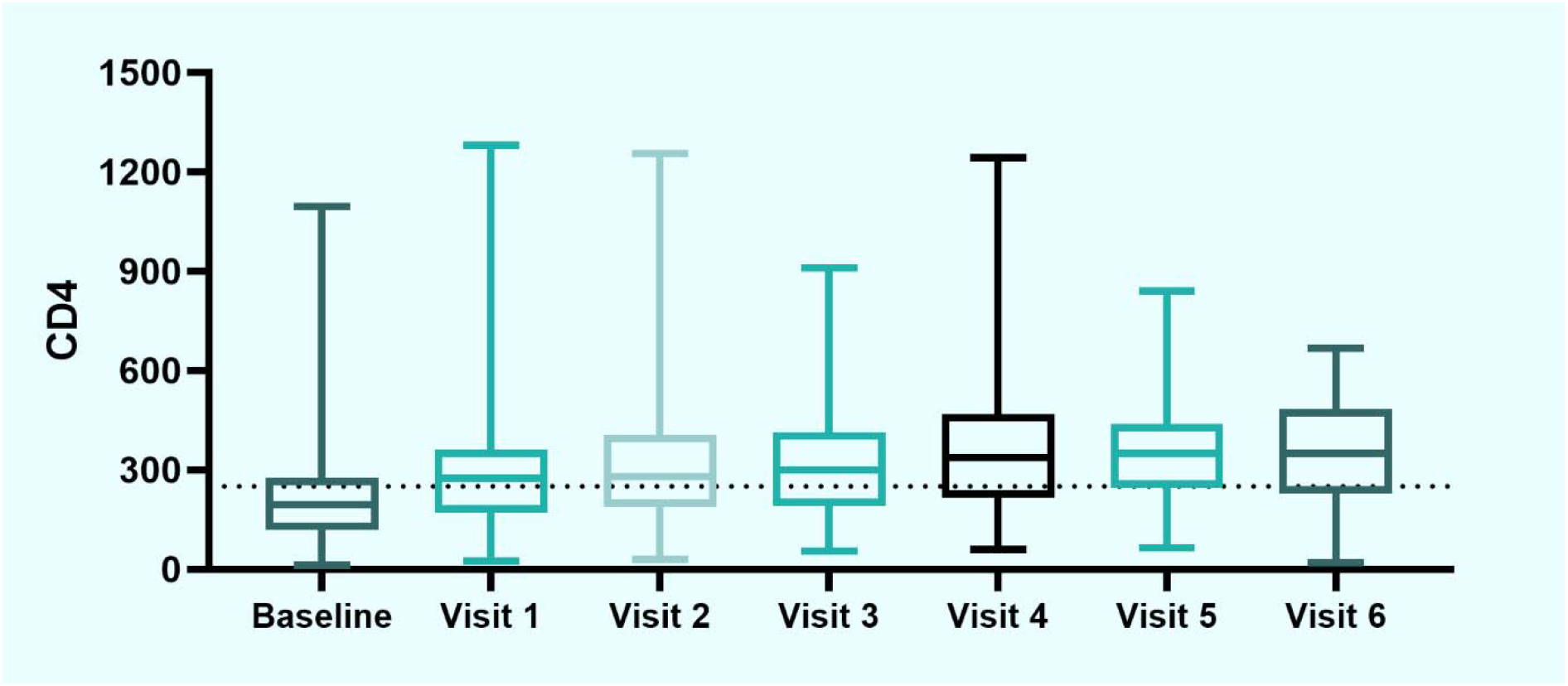
Variation in the median CD4 count of the participants from baseline through visit 6.

**Figure 2:**
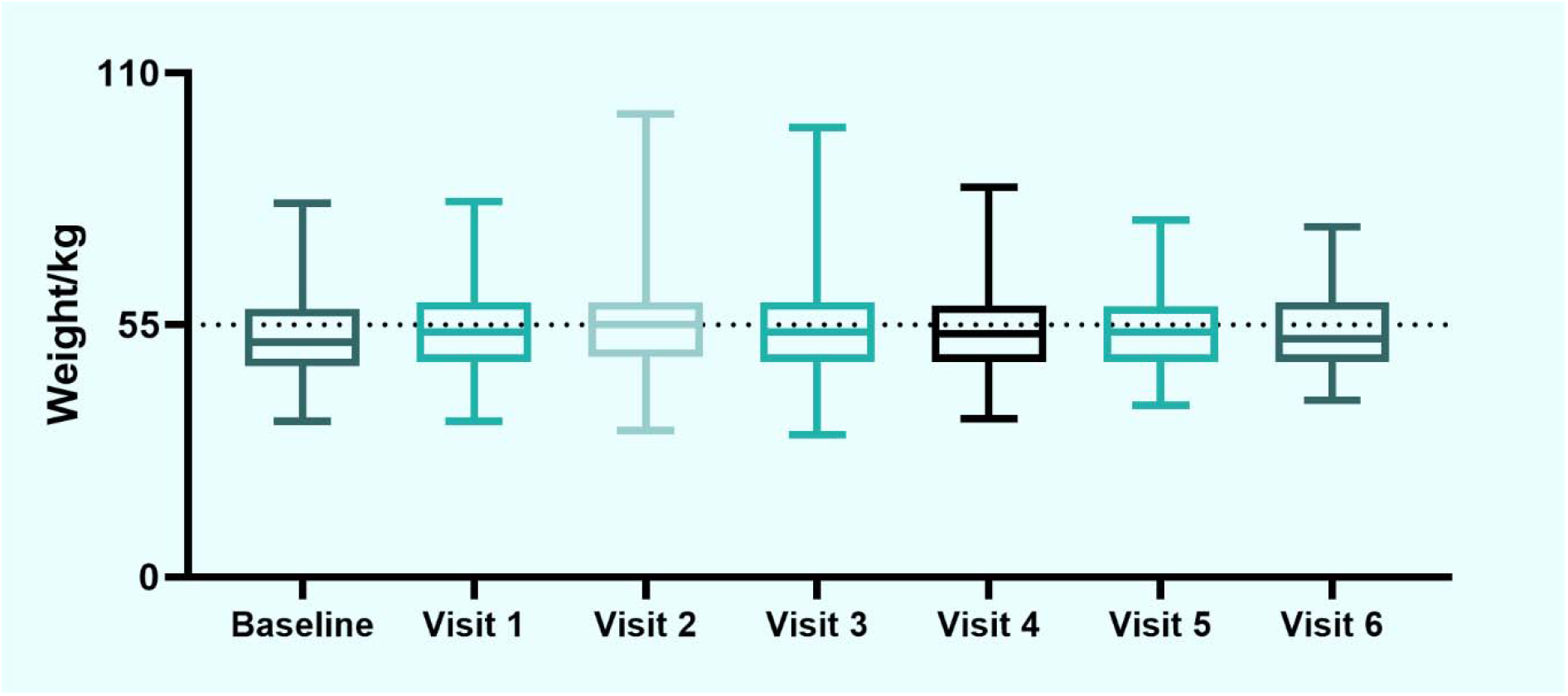
Variation in the median weight of the participants from the baseline through visit 6

Eighty-two (32.8%) patients changed their treatment regimen. Of these, 43(52.4%) had body fat abnormalities, 9(11%) had neuropathy, 1(1.2%) had treatment failure and the rest had other form of side effects like severe anemia (n=1), drug toxicity (n=1), Stevens-Johnson syndrome (n=1) and virological failure (n=1).

43(17.2%) of the patients had body fat abnormalities with a median(range) age of 35(20-63) years (p-0.6) and also the median(range) body weight of those patients at baseline was 46.5(41-52) kg (p-54) compared to 51.3 kg.

Patients who had body fat abnormalities had a relatively lower Hb count with a median(range) count of 11.95(8.3-14.7) at baseline compared to those who had no body fat abnormalities with a median(range) HGB count of 12.5(7.6-18.3) (p-0.048). Moreover, they had a relatively higher BUN count with a median(range) count of 11(6-31) at baseline compared to those who had no body fat abnormalities with a median(range) BUN count of 9(0.6-48) (p-0.042) (**Table 2**).

**Table 2:**
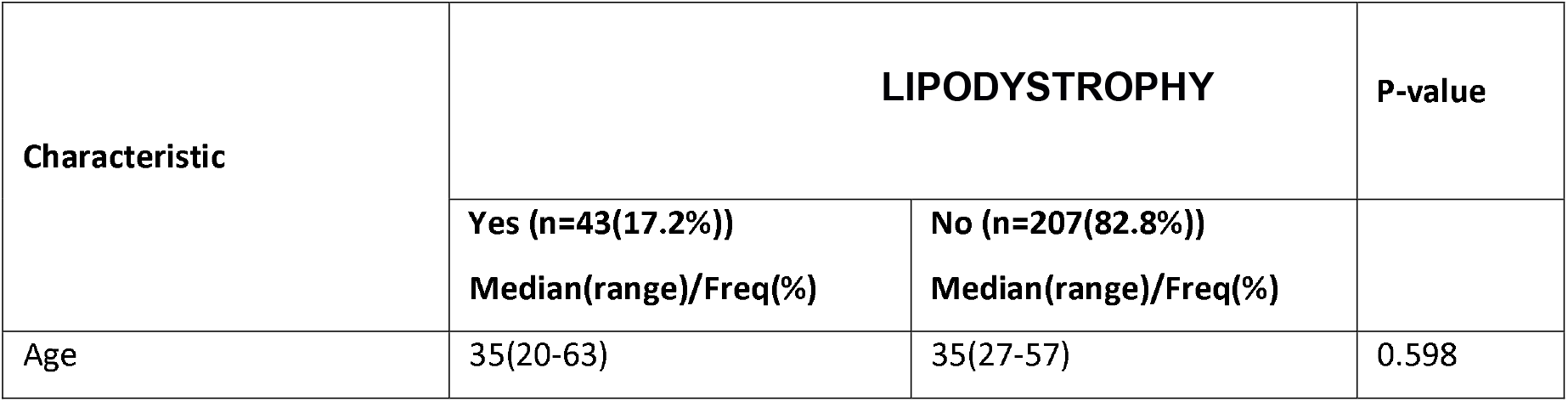
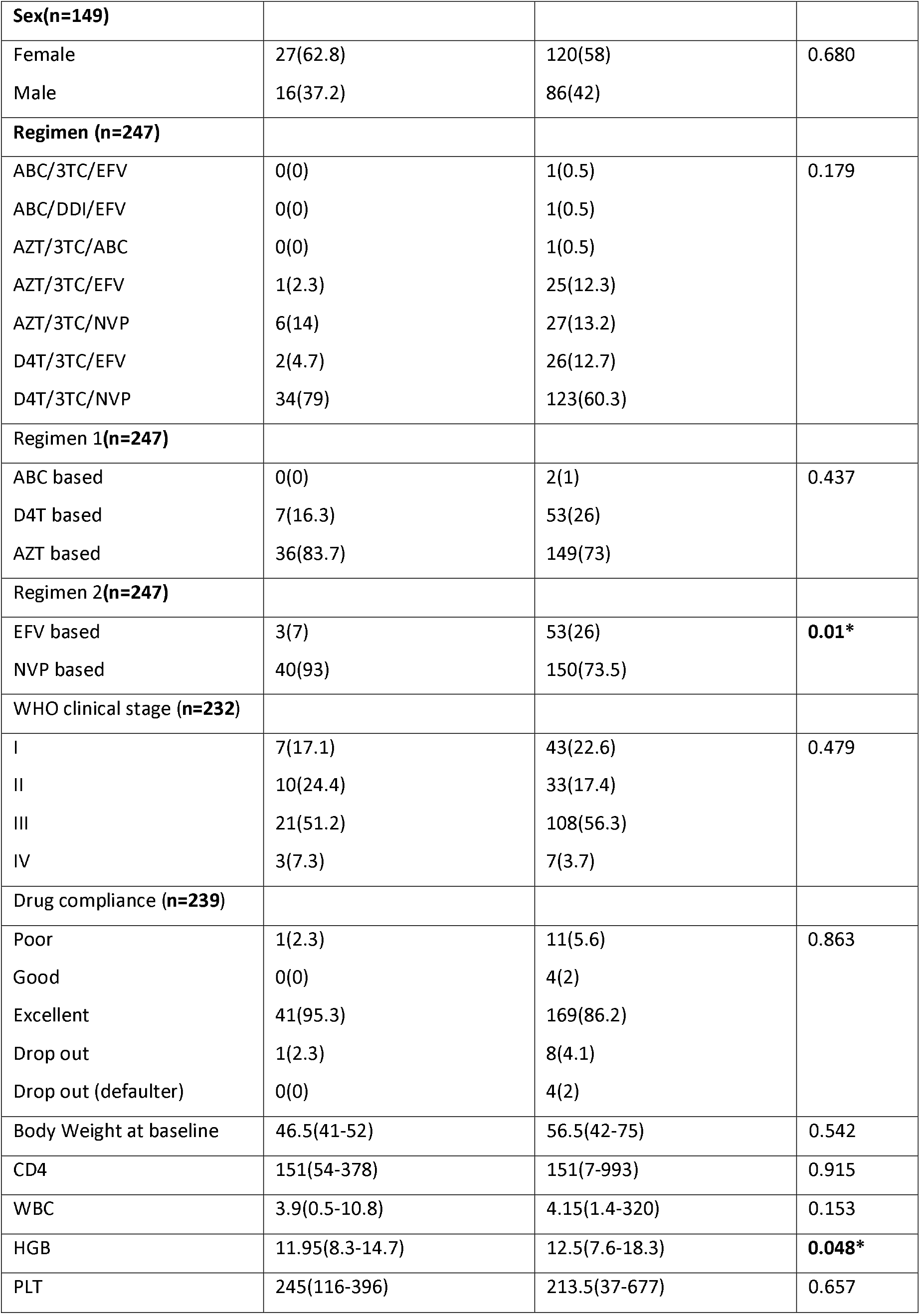
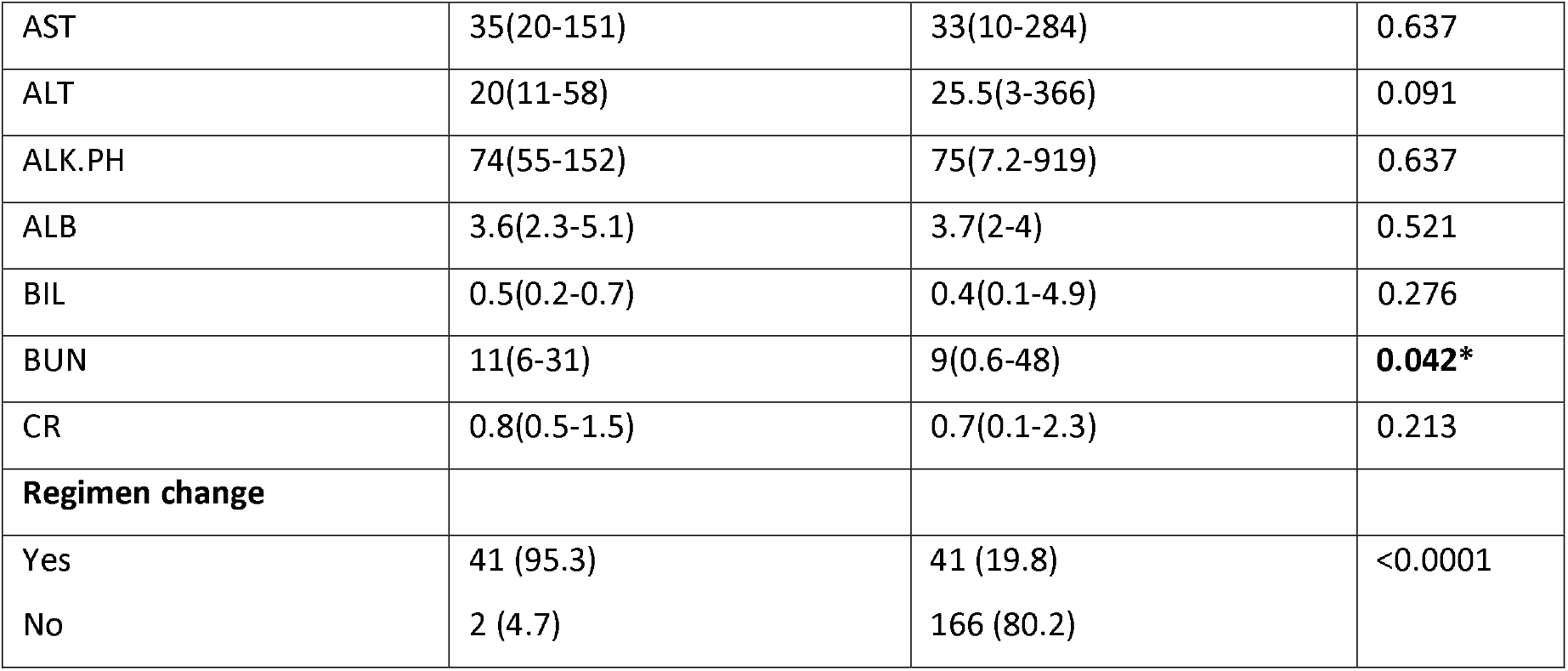
Sociodemographic and clinical factors associated lipodystrophy

Compared to NVP-based regimen, EFV-based regimen was significantly associated with lipodystrophy (p=0.01).

## Discussion

Body fat abnormalities in patients on ART is a real concern for patients and their relatives. The impact of body fat changes on patients affects drug adherence. This study was aimed at determining the prevalence of body fat abnormalities and associated factors in patients on ART at Orotta national referral hospital, Asmara Eritrea. No study has been done previously to address this abnormality in our setting.

The majority of participants were in stage III WHO clinical staging and had CD 4 count < 300 at the time of ART initiation. In one study increased age at initiation of ART have been associated with self-reported lipoatrophy but no specific age was determined (9). However, one study using objective measurements of lipoatrophy has not confirmed a link between age (14). The similar studies have also shown that, disease severity or high clinical staging at initiation of treatment is one of the predictors of fat atrophy (14,15). Hence in our study the severity of disease could have an input to the high percentage (17.2%) of body fat abnormalities.

The mean CD4 count at the initiation of therapy in this study was 151/mm^3.^ In one study, increased age and lower CD4 cell counts at initiation of HIV therapy have been associated with self-reported lipoatrophy which was consistent with our findings (9). The older the age at the start of ART and the earlier the development of body fat abnormalities seen in this study. Patients with the age of 50-59 years showed significant difference in developing body fat abnormalities when compared with other age groups. 50 percent from same age group developed side effect as early as 7 to 18 months of age. This demonstrated old age is one of the risk factors to develop drug side effect earlier than younger patients. In one study, patients were evaluated for signs of body fat abnormalities and the independent factors for moderate/severe lipoatrophy were increasing age, any use of stavudine, use of indinavir for longer than 2 years (9).

Peripheral and central lipoatrophy is an associated entity of HIV infection (16). After initiation of ART, the body fat maldistribution was also described with the drug regimens (17). However, the clinical significance of HIV associated lipohypertrophy came more slowly than lipoatrophy in studies. Some of the reasons of this delay in report were, the perception that fat loss is more readily noticeable than fat accumulation, patients tend to report fat loss than fat accumulation due to its negative cosmetic effects, and fat accumulation can be confounded with obesity of other causes (15,18).

ART-induced lipodystrophy which is characterized by peripheral and/or central fat loss or accumulation is described in patients on zidovudine, stavudine, didanosine and most protease inhibitors (19). The management of ART induced lipodystrophy is difficult and complex that includes switching of drug regimen to non lipodystrophic regimen, lifestyle modification, statins, metformin, fibrates, growth hormone and thiazolidinediones (20).

In our study, lipoatrophy held 50.6% of total side effects noted, while lipohypertrophy (‘buffalo hump’) held only 1.2%. d4T, 3TC, NVP results in 79 percent of lipoatrophy when compared to the side effects noted with the other regimen, AZT, 3TC, NVP resulted in 14 percent, d4T, 3TC, EFV in 4.7 percent and AZT, 3TC, EFV in 2.3 percent. Lamivudine (3TC) was constituent of all the four regimens. Stavudine (d4T), efavirenz (EFV), zidovudine (AZT), nevirapine (NVP) are each in two drug regimens. In vitro and in vivo studies suggest that thymidine analogues – zidovudine and stavudine in particular, efavirenz play an important role in pathogenesis of lipoatrophy (21,22). In one prospective, randomized, open trial, 237 patients initiating ART were assigned to abacavir or stavudine, both combined with lamivudine or efavirenz (23).

Buffalo hump occurred in one male patient who was on d4T, 3TC, NVP. The use of protease inhibitor therapy was initially thought to lead to the development of fat deposition (24). In a study done on ART-naive patients, there was no difference in the development of regional fat in any of the three treatment arms utilizing a PI, a NNRTI, or a PI plus NNRTI strategy (25). In this study we only got one patient with the disorder. Hence it is uneasy to attribute it to any drug or drug regimen.

ART-induced lipodystrophy is common in low-, low-middle income settings with considerable impact to the patients. These impacts includes increased risk of metabolic diseases, compromised quality of life and poor drug adherence (26). Increased prevalence of ART induced lipoatrophy, lipohypertrophy and mixed syndrome in short term and long term use of ART have been reported in the resource limited settings. The main explanation given to the rise of this burden more than in high-income countries, is the lack of better alternative of drug regimens which are easily accessible in high-income settings (26–28).

The switch of drug due to fat maldistribution was mainly from nevirapine to efavirenz or/and from stavudine to zidovudine. And drug regimen with efavirenz showed to be significantly associated with lipodystrophy (p-0.01). Our study observed that the huge burden of shifting drug regimen was due to lipoatrophy (more than half of the reason). In one randomized trial, limb fat loss was more common in subjects receiving NRTIs plus efavirenz compared to NRTIs plus the PI lopinavir/ritonavir, similar to our finding (29). Our study demonstrated an excellent adherence which was 85%. Adherence is a key to successful therapy. Patients’ adherence to ART is a dynamic process that interacts with adipose tissue advancement. Better adherence is associated with a higher risk of subsequent occurrence of fat maldistribution (30). Patients who are started on ART need to be counseled about the importance of adherence prior to initiation of medications.

In 2019, the WHO revised the first line ART regimens. The first line options from the new guidelines are **a)** TDF + 3TC (or FTC) + DTG, and as alternative regimen, **b)** TDF + 3TC + EFV, and in special circumstances **c)** AZT + 3TC + EFV. Moreover, in 2016 WHO recommended countries to discontinue d4T use in first-line regimens due to well-recognized metabolic toxicities (31,32).

## Study limitations and strengths

Due to its retrospect nature some investigations were missing, e.g For most patients’ lipid panels were missing.

Body shape changes were only determined subjectively by clinicians’ clinical assessment which is prone to interobserver variability. Objective quantification of fat by CT, MRI, and DEXA scanning for lipoatrophy and quantitative measurements of visceral adipose tissue by CT or MRI were not done.

The effect of abacavir plus lamivudine plus efavirenz, abacavir plus didanosine plus efavirenz, zidovudine plus lamivudine plus abacavir could not be equated because of inadequate number of subjects on the mentioned regimens. There was only one subject in each regimen.

Since the initiation of ART in the country no study was done to address the real problem of body fat abnormalities in patients on ART. This study proved that the problem is real.

## Conclusion

The prevalence of body fat abnormality in our setting was 17.2 percent. The evolvement of body fat abnormality was between 19 and 36 months after initiation of ART. Drug regimens that include efavirenz showed to be significantly associated with lipodystrophy. In the revised 2016 WHO ART drug regimens, d4T was removed but AZT remained from the thymidine analogue, also 3TC, NVP and EFV. Therefore, using this study as an entry point, we recommend more multicentric prospective studies to be done on the role of AZT, 3TC, EFV, NVP, abacavir in the development of body fat maldistribution.

## Data Availability

The information used and/or analyzed during this manuscript is available from the corresponding author on reasonable request.

## Abbreviations

3TC: Lamivudine
AIDS: Acquired Immunodeficiency Syndrome.
ABC: Abacavir
ART: Antiretroviral Therapy/ Treatment
Atv/r: Atazanvir/ritonavir
AZT: Zidovudine
BMI: Body Mass Index
CT: Computed Tomography
DEXA: Dual-Energy X-ray Absorptiometry
d4T: Stavudine
ddI: Didanosine
EFV: Efavirenz
ELISA: Enzyme-Linked ImmunoSorbent Assay.
FTC: Emtricitabine
HIV: Human Immunodeficiency Virus.
HAART: Highly Active Antiretroviral Therapy/ Treatment
LPV/r: Lopinavir/ritonavir
MoH: Ministry of Health
MACS: Multicenter AIDS Cohort Study
MRI: Magnetic Resonance Imaging.
NVP: Nevirapine
NRTIs: Nucleoside or nucleotide Reverse Transcriptase Inhibitor
NNRTI: Non-Nucleoside Reverse Transcriptase Inhibitor
PI: Protease Inhibitor
SJS: Stevens-Johnson Syndrome
WHO: world health organization

## Declarations

### Ethics approval and consent to participate

Approval for the study was given by the Orotta school of medicine and the Research ethics and Protocol review committee of the Eritrean ministry of health. Administrative clearance was sought from Orotta National Referral Hospital. Permission was obtained from the authorities of the Hospital. All the data collected from the records of the respondents was kept confidential. Consent to participate was not obtained from the patients because the study was based on anonymized patient records.

## Authors’ contribution

All authors made substantial contributions to conception and design, acquisition of data, or analysis and interpretation of data; took part in drafting the article or revising it critically for important intellectual content; agreed to submit to the current journal; gave final approval of the version to be published; and agree to be accountable for all aspects of the work.

## Acknowledgements

We would like to acknowledge, the staff of HIV clinic of Orotta National referral hospital for their active support in the data collection process.

